# Environmental Profiles and COVID-19 Mortality Risk: A Latent Class Analysis of Intensive Care Patients in the Early Pandemic

**DOI:** 10.1101/2025.07.20.25331858

**Authors:** Wei Wang, Danning Li, Lauren Zhang, Matthew Eskell, Bryce Clark, Anand Damodaran, Randeep Mullhi, Tonny Veenith, Yajing An, Fang G Smith, Hui Li

## Abstract

**Background:** Coronavirus disease 2019 (COVID-19) continues to strain intensive care units (ICUs), particularly during seasonal surges and in conjunction with other respiratory infections. While clinical and demographic mortality predictors are well-established, the impact of environmental conditions remains less understood, especially at the individual level. We therefore analysed subgroups of ICU patients based on environmental conditions and evaluate differences in mortality risk.

**Methods:** In this retrospective, multi-centre cohort study, we analysed data from 1,166 adults admitted with COVID-19 to three Birmingham (UK) ICUs between March 1, 2020, and February 28, 2021. Using latent class analysis (LCA), we grouped patients by ambient temperature, relative humidity, and wind speed preceding ICU admission. Associations between class membership and ICU mortality were assessed using multivariable logistic regression, adjusting for age, sex, ethnicity, deprivation, BMI, frailty, and ICNARC Physiology Score. Relative risk (RR) of mortality was compared across classes.

**Results:** Four latent classes were identified. Two showed significantly increased mortality risk: one with high temperature and low humidity (RR=1.62, 95% CI: 1.14-2.29) and another with low temperature and moderate-to-high wind speed (RR=1.47, 95% CI: 1.15-1.89), compared to the reference class. Environmental and patient characteristics, particularly relative humidity, female sex, BMI, ICNARC score, and frailty, demonstrated class-specific associations with mortality.

**Conclusions:** Environmental exposures prior to ICU admission contribute to mortality risk in critically ill COVID-19 patients. Incorporating these conditions into patient risk stratification could enhance clinical decision-making, particularly during public health emergencies.

## Introduction

The Coronavirus disease 2019 (COVID-19) pandemic overwhelmed healthcare systems worldwide, particularly intensive care units (ICUs) [1,2]. It has resulted in over 230,000 reported deaths in the United Kingdom alone by June 2025 [3]. Now endemic, COVID-19 continues to drive seasonal surges that burden ICUs; for example, spikes in ICU admissions during late 2023 were linked to the co-circulation of COVID-19 and other respiratory infections [4–6].

Numerous studies have identified clinical, physiological [7–9], demographic, and socioeconomic factors, such as age, ethnicity, and obesity, as predictors of COVID-19 mortality [10–12]. However, associations between these factors and outcomes like hospitalisation or death have not always been consistent [13,14].

In parallel, an expanding body of research suggests that environmental factors, including temperature, humidity, windspeed, and ultraviolet (UV) radiation, may influence both viral transmission dynamics and host susceptibility [15–26]. Environmental factors have also been implicated in disease severity, with some studies reporting associations with hospitalisation, ICU admission, and mortality at an aggregate level rather than using individual patient data [27–35]. However, these findings are also inconsistent.

Our objectives were therefore threefold: (1) to identify and characterise latent classes based on environmental factors; (2) to investigate the association between class membership and mortality; and (3) to assess whether environmental profiles offer additional explanatory value in understanding mortality risk among ICU patients with COVID-19.

## Methods

### Study Design and Sample Size

This was a retrospective cohort study using data collected from ICU patients admitted to three hospitals between March 1, 2020, and February 28, 2021, under the University Hospitals Birmingham (UHB) National Health Service Foundation Trust: Birmingham Heartlands Hospital (BHH), Good Hope Hospital (GHH), and the Queen Elizabeth Hospital Birmingham (QEH). Physicians diagnosed patients with COVID-19, which was subsequently confirmed with Medical Wire & Equipment (MWE) ViroCult nasal PCR tests.

In total, 1,180 patients were admitted to the ICU of these hospitals during this timeframe. Fourteen had incomplete medical information and were excluded from final analyses. Thus, this cohort netted 1,166 usable observations: 116 from BHH, 63 from GHH, and 987 from QEH (Figure 1). All the observations for the study were de-identified.

**Figure 1.**
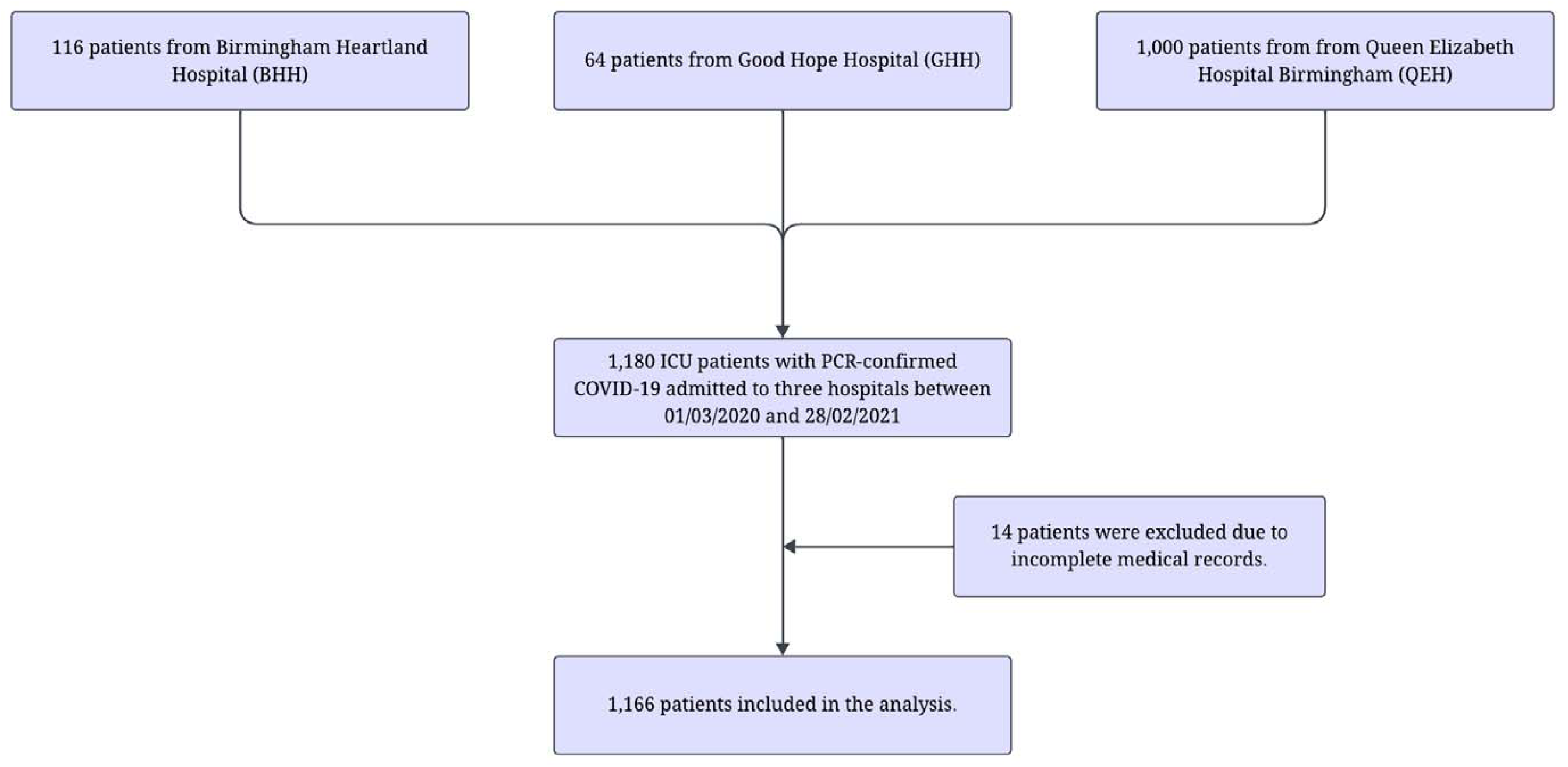
Study flow diagram.

### Data Collection

Data used in the study included both demographic and clinical factors. Demographic variables included age, sex, and ethnicity. Clinical data was collected from Intensive Care National Audit & Research Centre (ICNARC) by a data clerk and the UHB’s prescribing information and communication system (PICS System, Birmingham, England). These physiological variables included age, body mass index (BMI), frailty score, and ICNARC Physiology score (ICNARC score). Date of admission, ICNARC scores, and mortality status (dead/survival) at hospital discharge were taken from the ICNARC registry. Age and BMI were extracted from PICS.

The ICNARC score is an overall metric based on the weights in twelve physiological parameters during the first 24 hours following admission to the ICU. It includes heart rate, blood pressure, temperature, respiratory rate, oxygenation, arterial pH, serum creatinine, serum sodium, white blood cell count, Glasgow Coma Score, urine, and serum urea. It ranges from 0 to 100, with a higher score corresponding to increased severity[36].

The frailty score is a subjective estimate of an individual’s degree of frailty on a scale ranging from 1 to 9 (≥5 is considered frail). It was derived through a brief interview with the patient or family on their functional baseline and comorbidities conducted by the clerking doctor [37]. If there was no score documented in the notes for the patient’s index admission, the frailty score was allocated on review of the noting.

The Index of Multiple Deprivation (IMD) measures socioeconomic status [38–45]. Patients’ postcode data, extracted from ICNARC records, was matched to IMD deciles (1-10) using the Oxford Population Health tool, with 1 representing the most deprived 10% of areas in England and 10 the least deprived. An indicator variable, “deprived,” was constructed to represent individuals in the lowest deciles (IMD ≤ 2).

All environmental data during the study period were extracted from the <u>Visual Crossing database</u> (accessed on 15 June 2023). We selected three daily meteorological variables: temperature (°C), humidity (%), and windspeed (km/h) based on their relevance in shaping environmental viral persistence and potential effects on host susceptibility, as supported by prior literature. For each patient, environmental data were matched to the date of ICU admission and their residential postcode. Estimates of the COVID-19 incubation period, or the time between exposure to the virus and the onset of symptoms, range from 2 to 14 days [46,47]. To account for short-term fluctuations, the environmental variables were smoothed using a seven-day backward moving average, counted from the date of ICU admission.

### Blinding and Sample Size

The nature of the modelling and data handling meant that it was not possible to blind statisticians to outcomes. The authors were blinded when assessing the frailty score, as was the ICNARC data clerk when retrieving data. All patients admitted to the three ICUs from March 2020 to February 2021 were included to avoid sampling bias.

### Statistical Analysis

For this study, we used temperature, humidity, and windspeed as class-defining variables. To improve interpretability, these continuous variables were further discretised into sub-categories [48,49]. Temperature was stratified using 0°C as the reference point, in increments of 5°C. The final temperature classification consists of three categories: cold (<5°C), mild (5-10°C), and warm (>10°C). Due to paucity of extreme data points (temperatures below 0°C and above 15°C represented 0.09% and 4.2% of observations, respectively), these observations were grouped into adjacent categories. This empirically derived categorisation aligns with the thresholds in prior studies using similar cutoffs to capture nonlinear temperature effects on health outcomes [50,51] and follows best practices for combining sparse categories to enhance model stability [52].

Humidity was also categorised into three groups: dry (<70%), moderate (70-90%), and humid (>90%), based on data distribution and environmental health literature. Low humidity weakens innate immune defence and enhances virus transmission by prolonging aerosol stability and infectivity [53,54]. Conversely, high humidity may promote microbial growth indoors, contributing to respiratory inflammation risk [55,56].

Similarly, the variable windspeed was categorised into three levels: gentle (<17 km/h), windy (17–23 km/h), and strong (>23 km/h). These classifications were based on data distribution and established thresholds in environmental epidemiology. Studies suggest that lower wind speeds are linked to increased viral aerosol stability and reduced dispersion, potentially enhancing transmission [57,58].

We then employed LCA to identify distinct environmental profiles. Seven sequential models were fitted, ranging from one to seven classes. Model fit was evaluated using Akaike Information Criterion (AIC), Bayesian Information Criterion (BIC), consistent AIC (CAIC), and sample-size-adjusted BIC (SABIC) [59,60]. The Vuong-Lo-Mendell-Rubin adjusted likelihood ratio test (VLMR-LRT) was applied to compare models with k and k-1 classes, where a significant p-value indicates that the k-class model outperforms the k-1-class model [60,61].

Classification reliability was assessed using entropy and average latent class posterior probability (ALCPP) [62]. Entropy, ranging from 0 to 1, reflects class separation, with values ≥0.80 indicating reliable classification [63]. Similarly, ALCPP values ≥0.80 per class indicate confident assignment of individuals to their respective latent classes [60,64,65].

Multivariate logistic regression analyses were conducted separately within each latent class to evaluate associations of the covariates (clinical variables and environmental factors) with mortality, and results were reported as odds ratios (ORs). Relative risks (RRs) were calculated to compare mortality outcomes across latent classes. Continuous variables were standardised for comparability. To evaluate the relative contribution of covariates with differing units, we also calculated relative importance scores after normalising the data within each class [66]. Higher scores indicate a greater influence of the variable in defining class membership. All analyses were performed using R software (version 4.2.3).

## Results

### Data characteristics

In this cohort of 1,166 patients, 276 (23.7%) died during their ICU stay and 890 (76.3%) survived. Mortality rates varied by hospital, with 36.2% at BHH (42/116), 28.6% at GHH (18/63), and 21.9% at QEH (216/987). Descriptive statistics and variable definitions are presented in Table 1.

**Table 1.** Descriptive statistics.

| Variables | Description | Mean | Std Dev | Median | t-test/Chi-squared test |
| --- | --- | --- | --- | --- | --- |
| Status | Indicator variable of mortality: death=1, survival=0 | 0.24 | 0.43 | 0 | <0.001 |
| Age | Age of a patient in years | 57.25 | 13.22 | 58.00 | <0.001 |
| Female | An indicator variable: female=1, male=0 | 0.33 | 0.47 | 0 | 0.168 |
| IMD | Index of Multiple Deprivation: ranging from 1 to 10, with 1 referring to the most deprived and 10 the least deprived areas | 3.25 | 2.75 | 2.00 | 0.216 |
| Deprived | Living in deprived area: if IMD equals 1 or 2 then deprived =1; else 0 | 0.57 | 0.50 | 1.00 | 0.268 |
| BMI | Body Mass Index: body mass divided by the square of the body height, in units of kg/m<sup>2</sup> | 30.93 | 6.76 | 29.39 | <0.001 |
| Frailty score | An estimate of an individual's degree of frailty on a scale of 1 (very fit) to 9 (terminally ill) | 2.76 | 0.87 | 3.00 | <0.001 |
| ICNARC score | A measure of severity of illness, with an integer score between 0 and 100 based on weights in 12 physiological parameters during the first 24 hours following admission to the ICU; higher score correlates to increased severity. | 21.30 | 15.19 | 18.00 | <0.001 |
| Temperature | Average daily temperature in degrees Celsius (7-day moving average) | 6.86 | 4.04 | 6.07 | 0.039 |
| Humidity | The amount of water vapour present in the air compared the maximum amount possible for a given temperature, expressed as a mean percentage (7-day moving average) | 82.06 | 10.33 | 86.34 | <0.001 |
| Windspeed | The two-minute average of wind speed in km/hour (7-day moving average) | 19.74 | 3.74 | 19.51 | 0.005 |
| White | Indicator variable: White=1, 0 otherwise. | 0.45 | 0.50 | 0 | 0.238 |
| Asian | Indicator variable, Asian=1, 0 otherwise. | 0.34 | 0.48 | 0 | 0.960 |
| Black | Indicator variable, Black=1, 0 otherwise. | 0.05 | 0.22 | 0 | 0.267 |
| Mixed/others | Indicator variable, mixed or other ethnicities=1, 0 otherwise. | 0.08 | 0.27 | 0 | 0.098 |
| Unknown | Indicator variable, unknown ethnicity=1, 0 otherwise. | 0.07 | 0.25 | 0 | 0.089 |

The cohort had a mean age of 57.3 (SD=13.2) years, with 384 (33%) female patients. 45% were White and 34% were Asian. 57% of patients were from deprived areas, with a median IMD of 2. Half of the patients were overweight with a median BMI of 29.39 and mean BMI of 30.93 (SD=6.76). The patients also had a mean frailty score of 2.76 (SD=0.87) and ICNARC score of 21.3 (SD=15.2).

During the study period, patients were admitted to ICUs under weather conditions with an average temperature of 6.9°C, relative humidity of 82.1%, and wind speed of 19.7 km/h. The median temperature was 6.1°C, relative humidity was 86.3%, and wind speed was 19.5 km/h.

Figure 2 superimposes weekly counts of ICU survivals and deaths over the 52-week period with background environmental data. Two peaks in mortality and survival were observed at weeks 4-6 and weeks 44-47. Temperature increased gradually until week 24, then declined into winter weeks. Humidity dropped sharply in the first seven weeks, fluctuated moderately until week 30, and remained high from week 31 onward. Wind speed was stable until week 25, then exhibited moderate fluctuations particularly in the later weeks.

**Figure 2.**
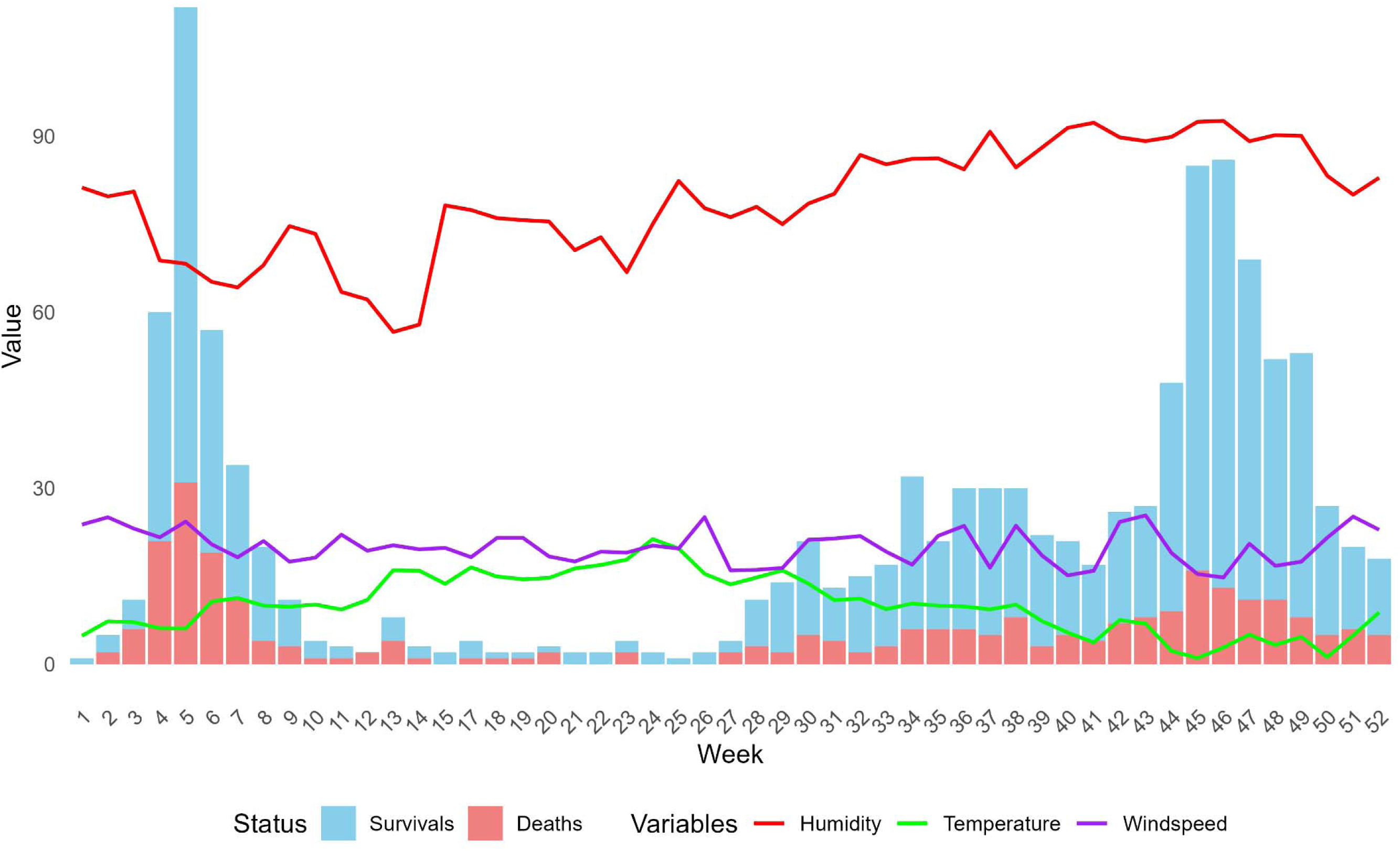
Weekly variations in mortality outcomes and environmental variables.

**Figure 3.**
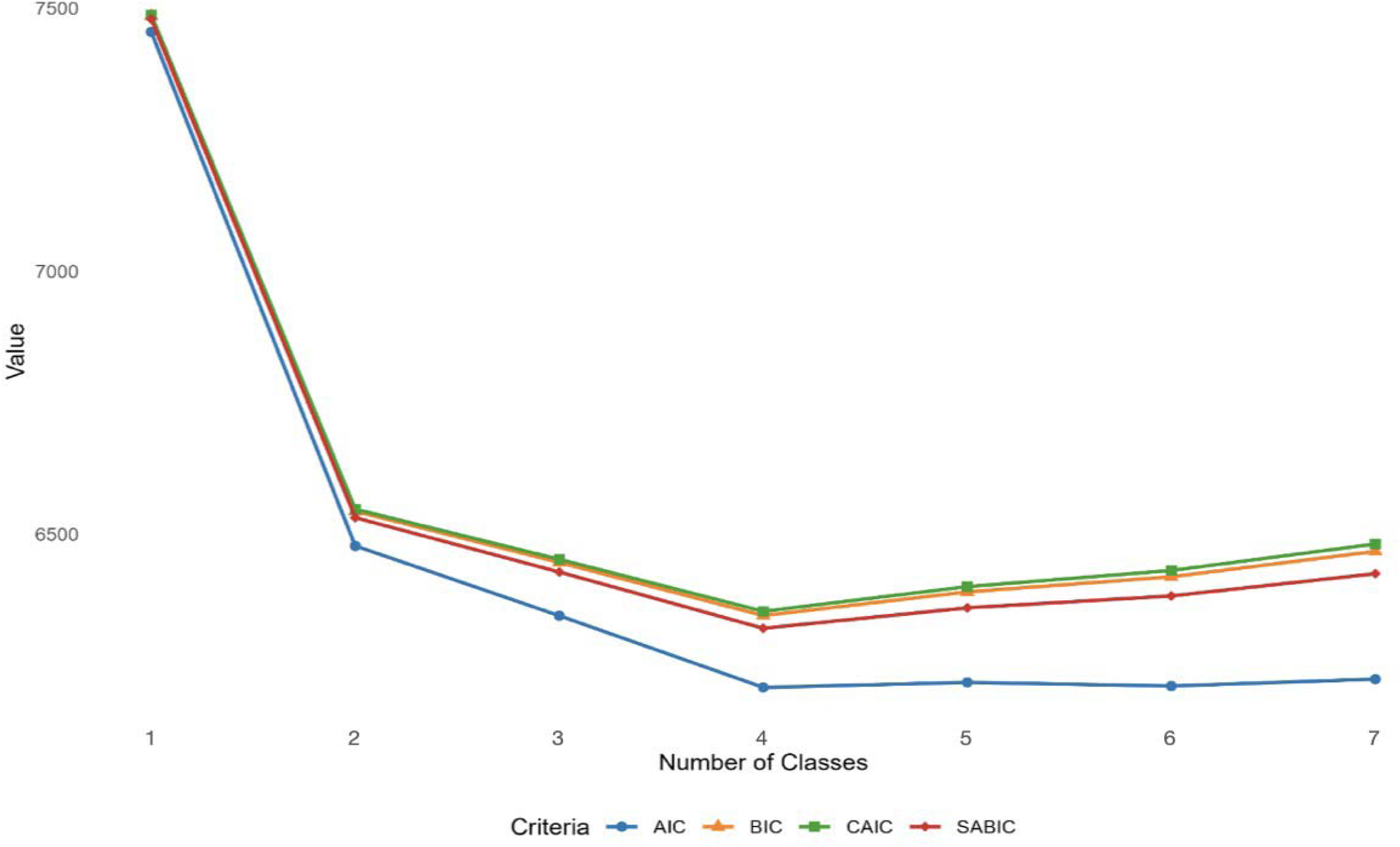
Model fit criteria across number of classes.

### Characteristics of LCA-defined patients

Model selection favoured the four-class solution, which had the lowest AIC, BIC, CAIC, and SABIC values (Figure 2), indicating optimal fit. Fit indices improved with additional classes up to four, after which they worsened. The VLMR-LRT supported this, with significant p-values (p<0.01) up to four classes and a non-significant result (p=0.724) for the five-class model. Though not used for selection, entropy was high (0.904), and ALCPP values exceeded 0.80 (lowest: 0.859), indicating strong classification quality. Model diagnostics are summarized in Table 2.

**Table 2.**
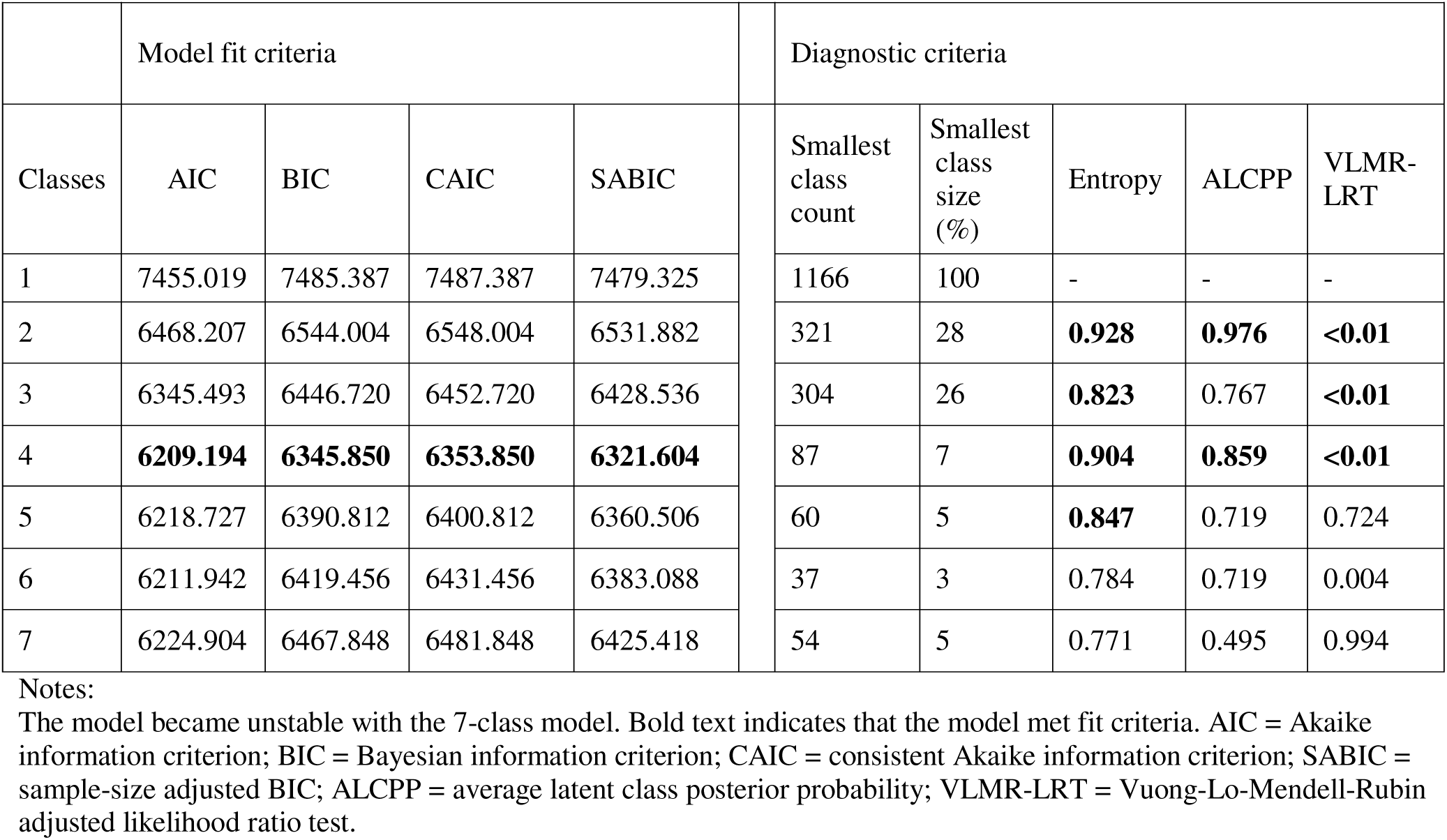
Summary of latent class model evaluation.

### Characteristics of environmental profiles

The posterior probabilities, or likelihood of class membership based on environmental variables, were calculated for each of the four latent classes and illustrated in Figure 4 and Supplemental material Table 1.

**Figure 4.**
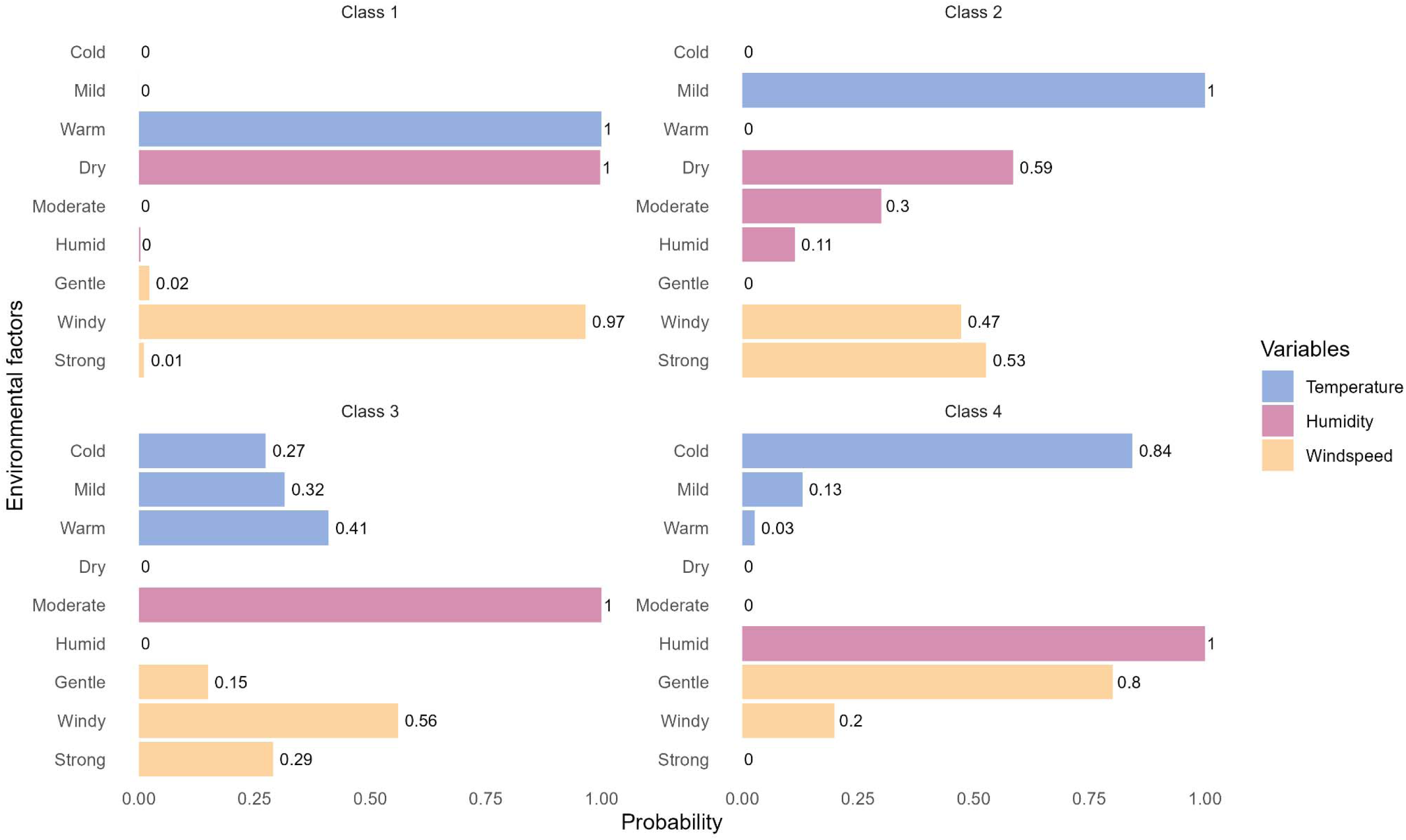
Environmental profiles across classes. On the graph, the horizontal axis represents the probability (or relative weight) of each environmental level within a class, while the vertical axis denotes the nine category levels: cold, mild, and warm (temperature); dry, moderate, and humid (humidity); and gentle, windy, and strong (wind speed). The three environmental variables are differentiated by a colour scheme: temperature in blue, humidity in pink, and wind speed in yellow. For example, Class 2 exhibited 100% probability of mild temperature, 59% likelihood of dry conditions, and 0% probability of gentle wind.

**Class 1 (warm, dry, windy):** This was the smallest group with 87 patients (7% of the sample) defined by warm temperatures (100% probability), dry conditions (100% probability), and predominantly windy conditions (97% probability). Patients in this class experienced consistently warm, dry environments with minimal wind speed variation. The high posterior probabilities across all environmental variables suggest a highly homogeneous profile for Class 1.

**Class 2 (mild temperature):** This class, comprising 304 patients (26% of the sample), was characterised by mild temperatures (100% probability), and varied patterns of humidity and wind speed. 59% of patients experienced dry, 30% moderate, and 11% humid conditions. Wind speed was similarly heterogeneous, with 47% experiencing windy conditions and 53% experiencing strong winds.

**Class 3 (moderate humidity):** This was the largest class comprising 404 patients (39% of the sample) and was defined by moderate humidity (100% probability). Temperature conditions varied across the class, with 27% of patients experiencing cold, 32% mild, and 41% warm temperatures. Wind speed was similarly diverse: 56% experienced windy conditions, 29% strong wind, and 15% gentle breeze.

**Class 4 (cold, humid, gentle):** This class included 303 patients (28% of the sample) and was characterised mainly by humid weather (100% probability), cold temperatures (84% probability), and a gentle breeze (80% probability). A small proportion of individuals experienced mild temperatures (13%) or windy conditions (20%), while warm temperatures (3%) and strong winds (0%) were negligible.

### Time Trends in ICU Admissions

Time trends in ICU admissions varied distinctly across environmental classes (Figure 5). Class 1 (warm, dry, windy) exhibited a sharp peak in early spring (weeks 1-6), with minimal admissions thereafter, spanning 11 weeks. Class 2 (mild temperature) showed a bimodal distribution over 18 weeks, with a primary peak in late winter (weeks 6-10) and a secondary peak in early autumn (weeks 40-45). Class 3 (moderate humidity) had a broad, sustained distribution across 35 weeks, with admissions throughout the latter half of the year (weeks 20-52), peaking at weeks 35-40 and 45-50. Conversely, Class 4 (cold, humid, gentle) displayed a concentrated peak in late autumn and early winter (weeks 40-52), spanning 10 weeks.

**Figure 5.**
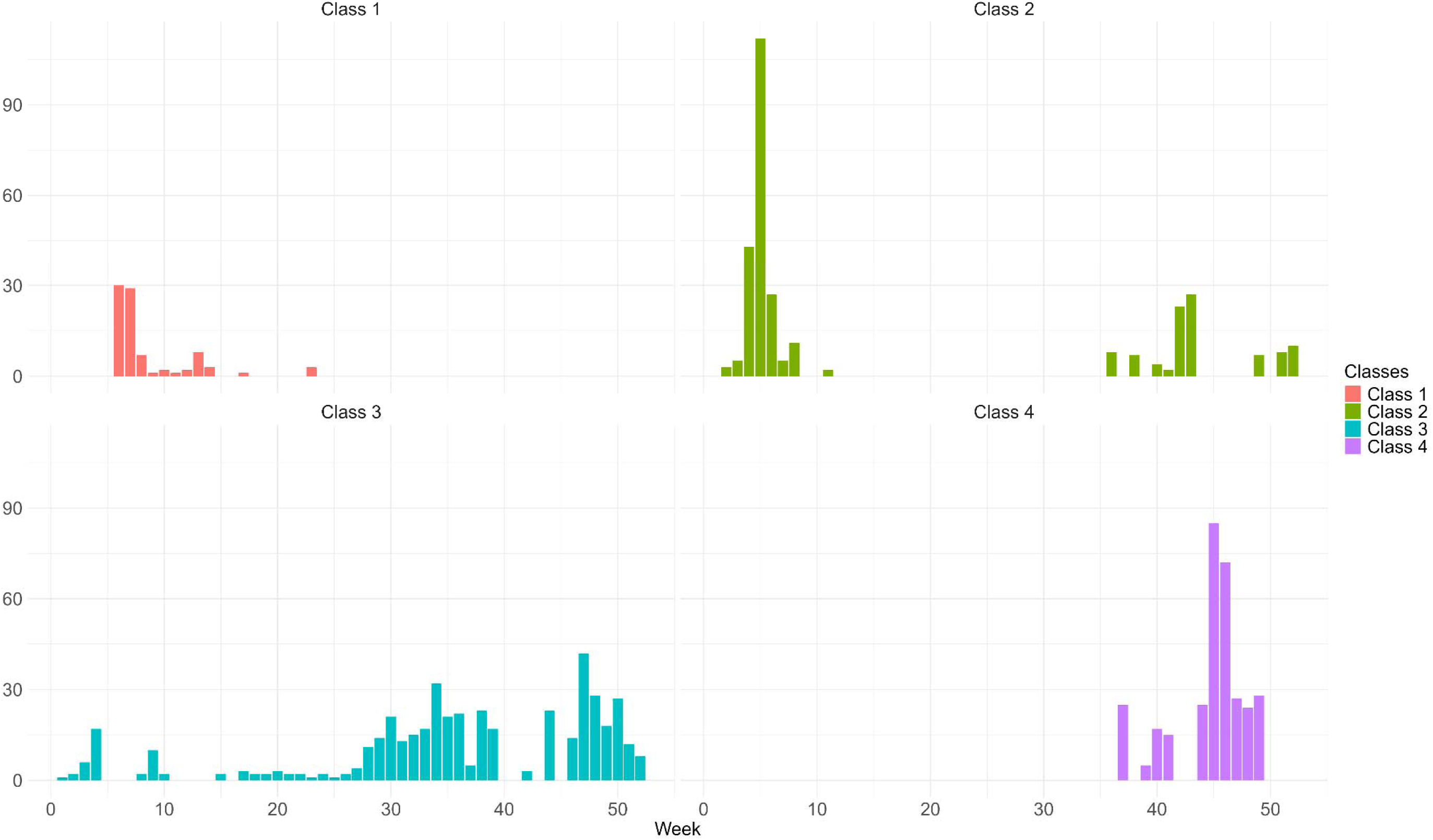
Weekly distribution of ICU admissions across classes.

### Mean comparison of class characteristics

The comparison of demographic, socioeconomic, and clinical characteristics across the four latent classes is summarised in Table 3. Differences in age, sex (female), ethnicity (White), and deprivation status were not statistically significant based on Kruskal-Wallis and Pearson Chi-squared tests. Although the overall distribution of ethnicity (White) was not significantly different (p=0.162), pairwise comparison between Class 2 and Class 3 indicated a marginal difference (p=0.078).

**Table 3.** Summary of patient characteristics and class comparisons.

|  | Class 1<br>Mean | Class 2<br>Mean | Class 3<br>Mean | Class 4<br>Mean | Class 1 vs<br>Class 2 | Class 1 vs<br>Class 3 | Class 1 vs<br>Class 4 | Class 2 vs<br>Class 3 | Class 2 vs<br>Class 4 | Class 3 vs<br>Class 4 | Kruskal<br>Wallis/Chi<br>-squared |
| --- | --- | --- | --- | --- | --- | --- | --- | --- | --- | --- | --- |
| Age | 58.563 | 56.914 | 56.752 | 57.916 | 0.287 | 0.233 | 0.675 | 0.868 | 0.326 | 0.230 | 0.438 |
| Female | 0.298 | 0.296 | 0.354 | 0.334 | 1.000 | 0.386 | 0.618 | 0.114 | 0.344 | 0.625 | 0.366 |
| White | 0.517 | 0.484 | 0.416 | 0.458 | 0.665 | 0.103 | 0.391 | **0.078**<sup>^</sup> | 0.579 | 0.273 | 0.162 |
| Deprived | 0.504 | 0.546 | 0.577 | 0.579 | 1.000 | 0.600 | 0.599 | 0.436 | 0.454 | 1.000 | 0.757 |
| BMI | 29.719 | 30.819 | 30.931 | 31.357 | 0.156 | 0.107 | **0.034** * | 0.825 | 0.311 | 0.389 | **0.089**<sup>^</sup> |
| Frailty score | 2.724 | 2.641 | 2.796 | 2.824 | 0.672 | **0.045** * | **0.018** * | **0.016** * | **0.005** ** | 0.566 | **0.007** ** |
| ICNARC<br>score | 24.695 | 22.610 | 19.812 | 21.245 | 0.279 | **0.001** ** | **0.070**<sup>^</sup> | **0.014** * | 0.246 | 0.187 | **<0.001** *** |
| Temperature | 12.494 | 7.055 | 8.232 | 3.256 | **<0.001** *** | **<0.001** *** | **<0.001** *** | **<0.001** *** | **<0.001** *** | **<0.001** *** | **<0.001** *** |
| Humidity | 63.587 | 74.115 | 83.931 | 92.027 | **<0.001** *** | **<0.001** *** | **<0.001** *** | **<0.001** *** | **<0.001** *** | **<0.001** *** | **<0.001** *** |
| Windspeed | 18.968 | 23.311 | 20.437 | 15.616 | **<0.001** *** | **<0.001** *** | **<0.001** *** | **<0.001** *** | **<0.001** *** | **<0.001** *** | **<0.001** *** |
| Sample size | 87 | 304 | 452 | 323 |  |  |  |  |  |  |  |

Notes: (1) \*\*\*, \*\*, \* and <sup>^</sup> are used for significance levels of 0.001, 0.01, 0.05 and 0.1, respectively. (2) p-values were reported for class comparisons. The Kruskal-Wallis test was used and the p-value reported for the continuous variables age, BMI, ICNARC score, temperature, windspeed, and humidity. For all other categorical variables, Pearson's chi-squared test or Mann-Whitney U test was conducted, and the p-value was reported.

Both frailty and ICNARC scores differed significantly across classes, with p-values of 0.007 and <0.001, respectively. Several pairwise comparisons also revealed significant differences, indicating substantial heterogeneity in clinical severity across the classes. While overall difference in BMI approached marginal significance at the 10% level (p=0.089), the pairwise comparison between Class 1 and Class 4 was statistically significant (p=0.03).

The standardised mean profiles across classes are illustrated in Figure 6. Minimal differences were observed in demographic and socioeconomic variables (age, sex, ethnicity, and deprivation), with standardised values remaining close to the overall sample mean. However, clinical variables, such as frailty and ICNARC scores, showed increased variation, with Class 1 exhibiting elevated levels of clinical severity. The most pronounced differences were in the environmental variables as they were used to define the latent classes. Notably, Classes 1 and 4 presented nearly opposing profiles across the environmental dimensions, while Classes 2 and 4 also showed contrasting patterns.

**Figure 6.**
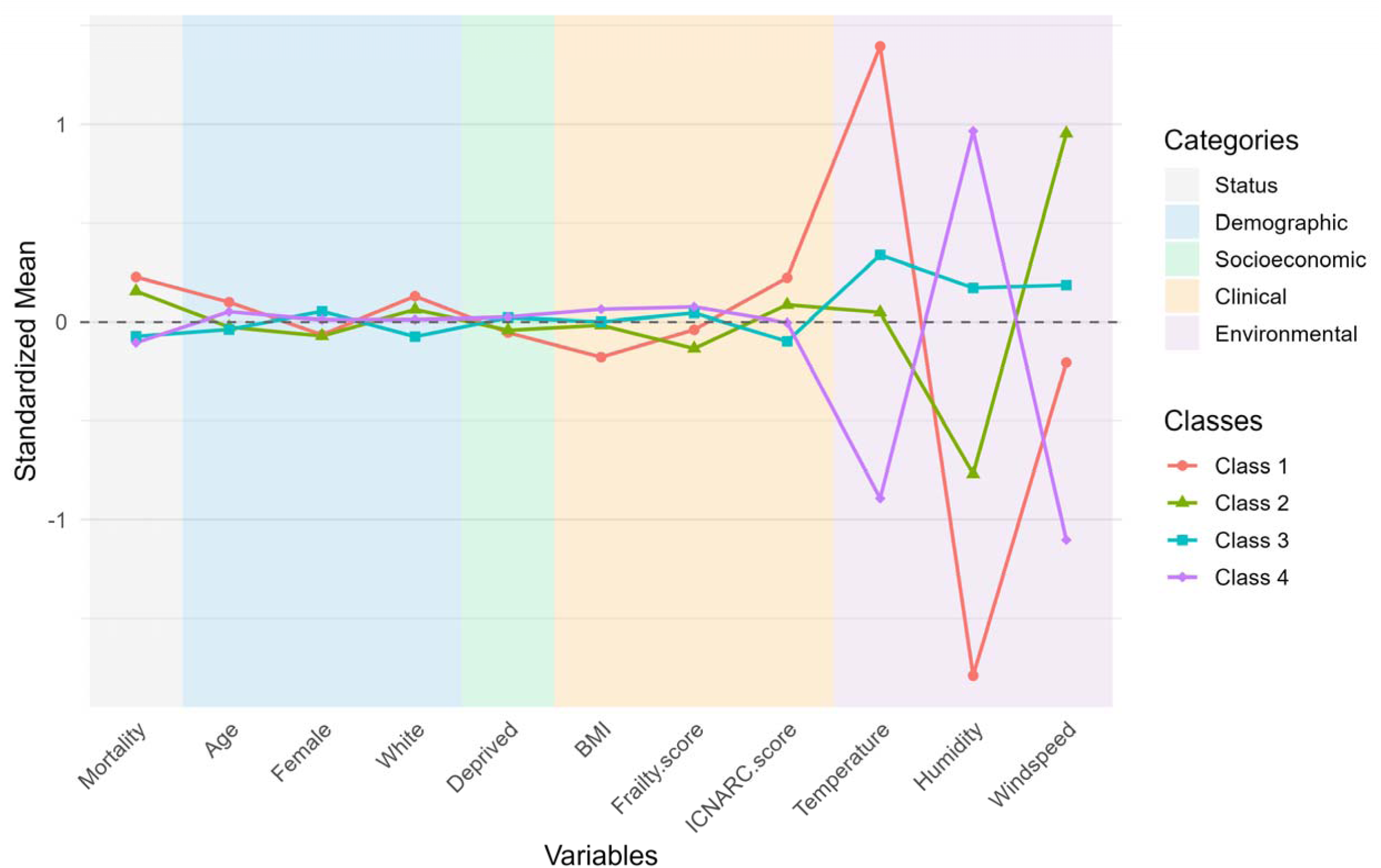
Standardised mean (Z-score) profiles of demographic, socioeconomic, clinical, and environmental variables across classes.

### Association of covariates with mortality within classes

#### Logistic Regression Results

Results from the multivariate logistic regression are given in Table 4, with ORs by class shown in Figure 7. In Class 2, older age (OR=1.843, p<0.001) and higher ICNARC scores (OR=1.630, p<0.001) were associated with increased mortality. In Class 3, mortality was linked to older age (OR=1.916, p<0.001), higher frailty scores (OR=1.340, p=0.023), and higher ICNARC scores (OR=1.667, p<0.01), while higher BMI (OR=0.711, p=0.028) and increased humidity (OR=0.520, p=0.033) were associated with lower mortality. In Class 4, female sex was associated with reduced mortality (OR=0.400, p=0.018), while older age, higher frailty scores, and higher ICNARC scores remained associated with increased mortality.

**Figure 7.**
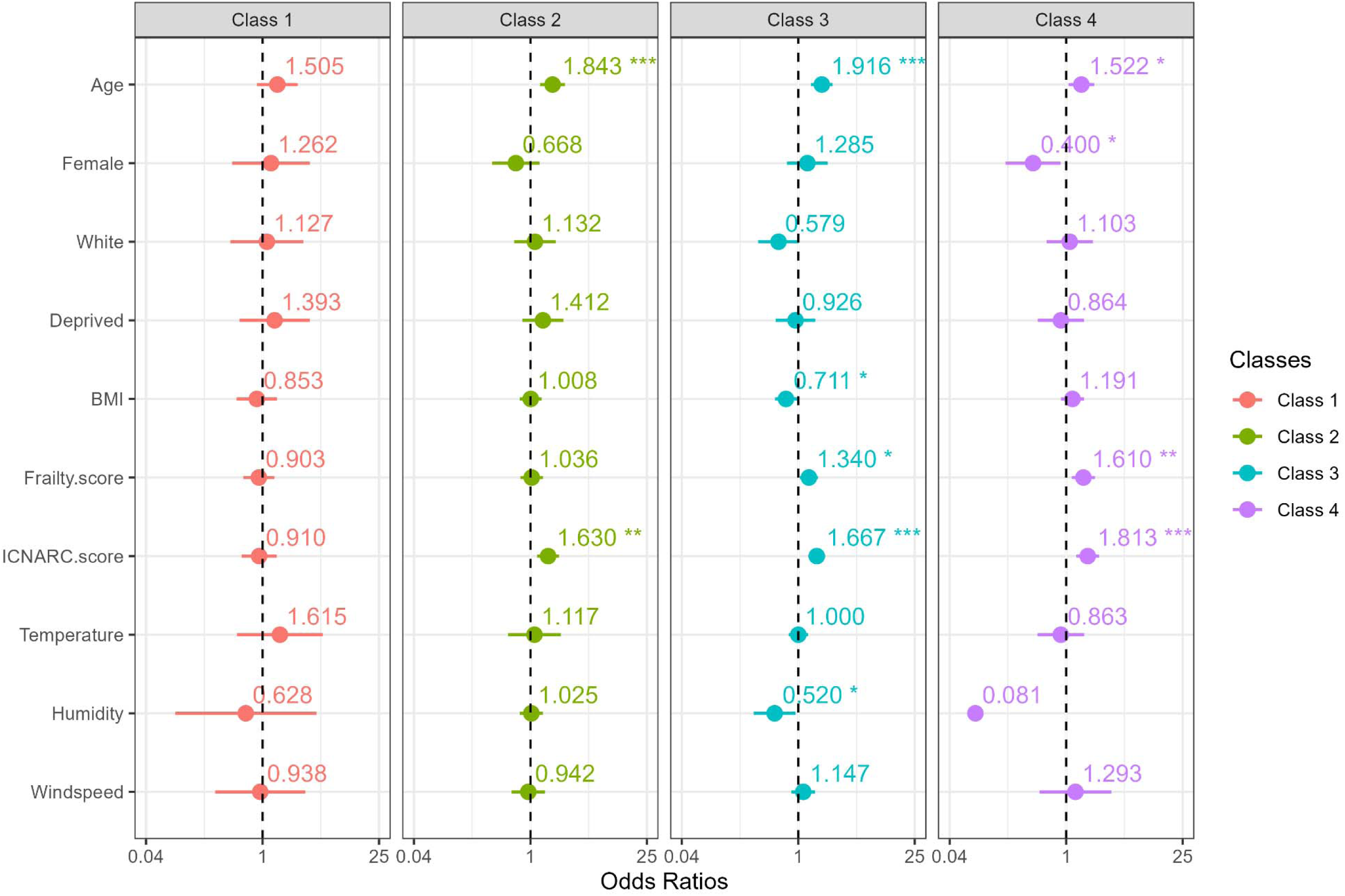
Odds ratios from multivariate logistic regression by class. Odds ratios and 95% confidence intervals for covariates associated with mortality stratified by latent class. Each panel represents one class, using a consistent colour scheme: Class 1 in red, Class 2 in green, Class 3 in blue, and Class 4 in purple.

**Table 4.**
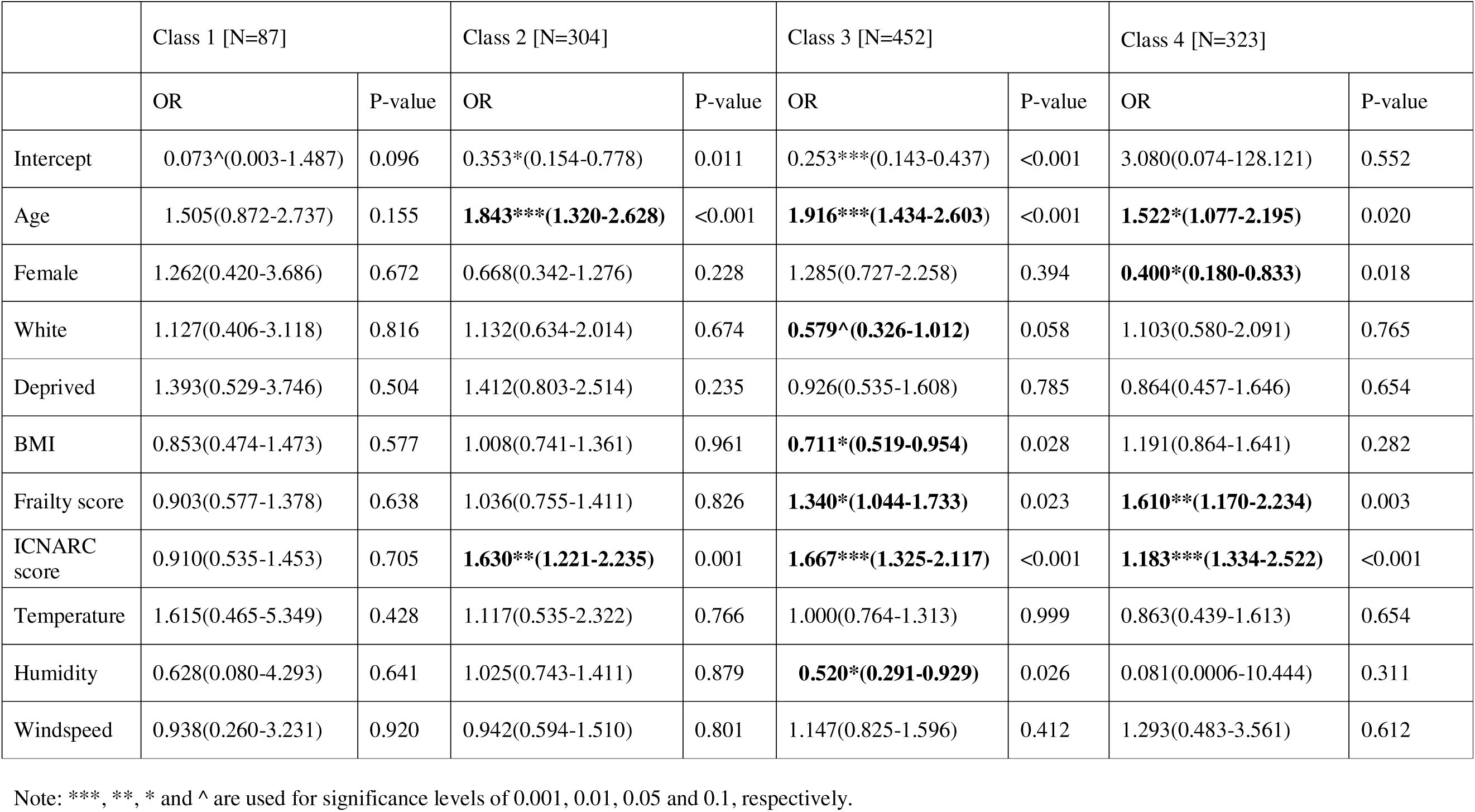
Results from multivariate logistic regression.

##### Relative Importance

There was substantial variation in the relative importance of variables across classes (Figure 8). Age emerged as a consistently important variable across all classes, with the highest relative importance observed in Class 3 (4.29). Similarly, the ICNARC score demonstrated high importance in Classes 2, 3, and 4 (3.82, 4.31, and 3.68, respectively). The frailty score was also notable in Classes 3 and 4. In contrast, variables such as windspeed and White ethnicity showed generally low importance across most classes; however, being of White ethnicity had a marked influence in Class 3 (1.90). When analysed by class, each latent subgroup exhibited a unique profile of dominant predictors. Class 1 was characterised by the relatively higher importance of age and temperature; Class 2 was characterised by higher importance of age and ICNARC score; Class 3 assigned higher importance to age, ICNARC score, frailty score, and humidity but lowest importance to living in a deprived area; and in Class 4, the frailty score, female sex, and ICNARC score were the most influential covariates.

**Figure 8.**
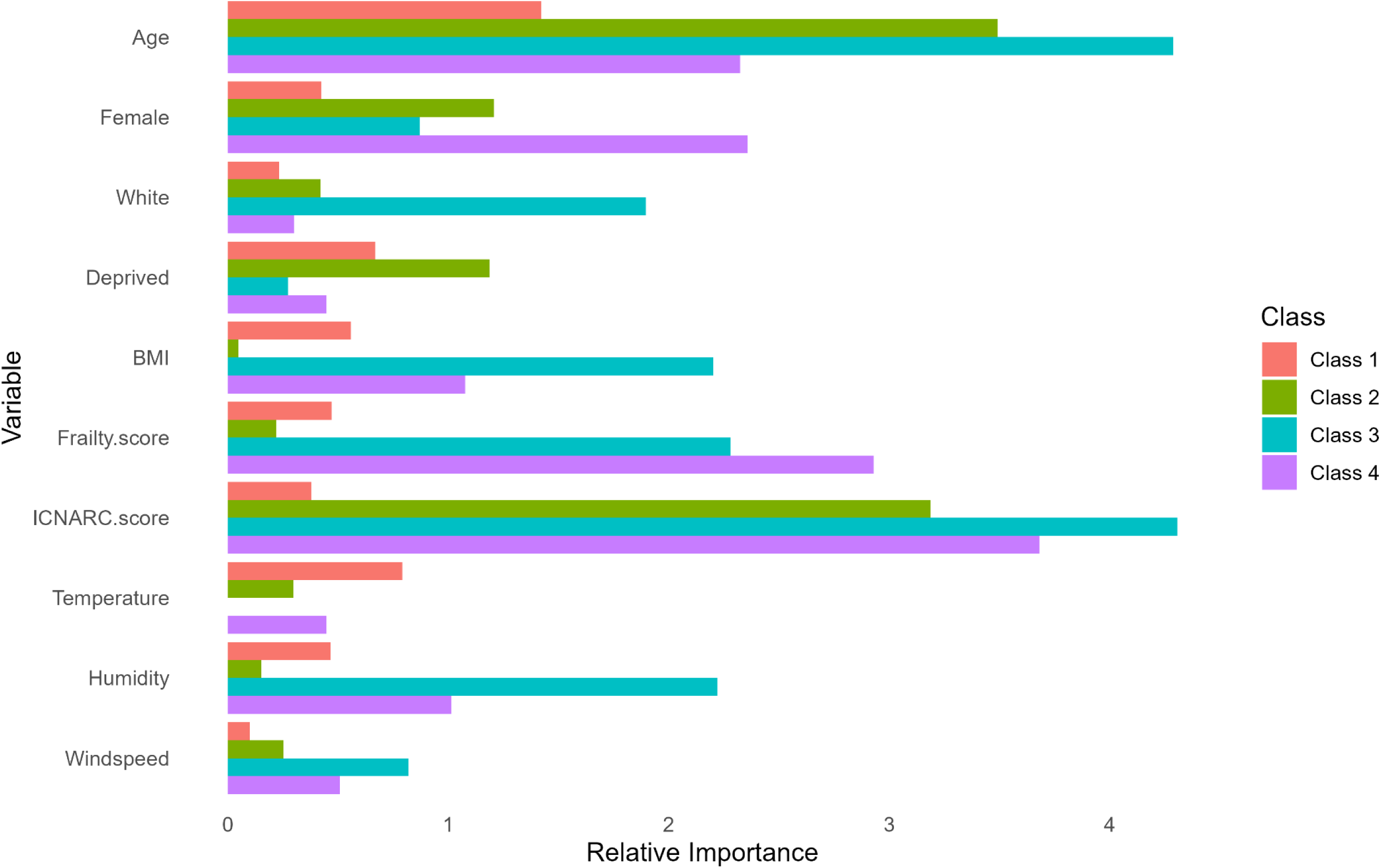
Relative importance scores of covariates across classes. The x-axis represents the relative importance score for each variable, quantifying its contribution to the predictive model for each class. The y-axis lists the covariates included in the analysis, and bars are colour-coded by latent class.

**Figure 9.**
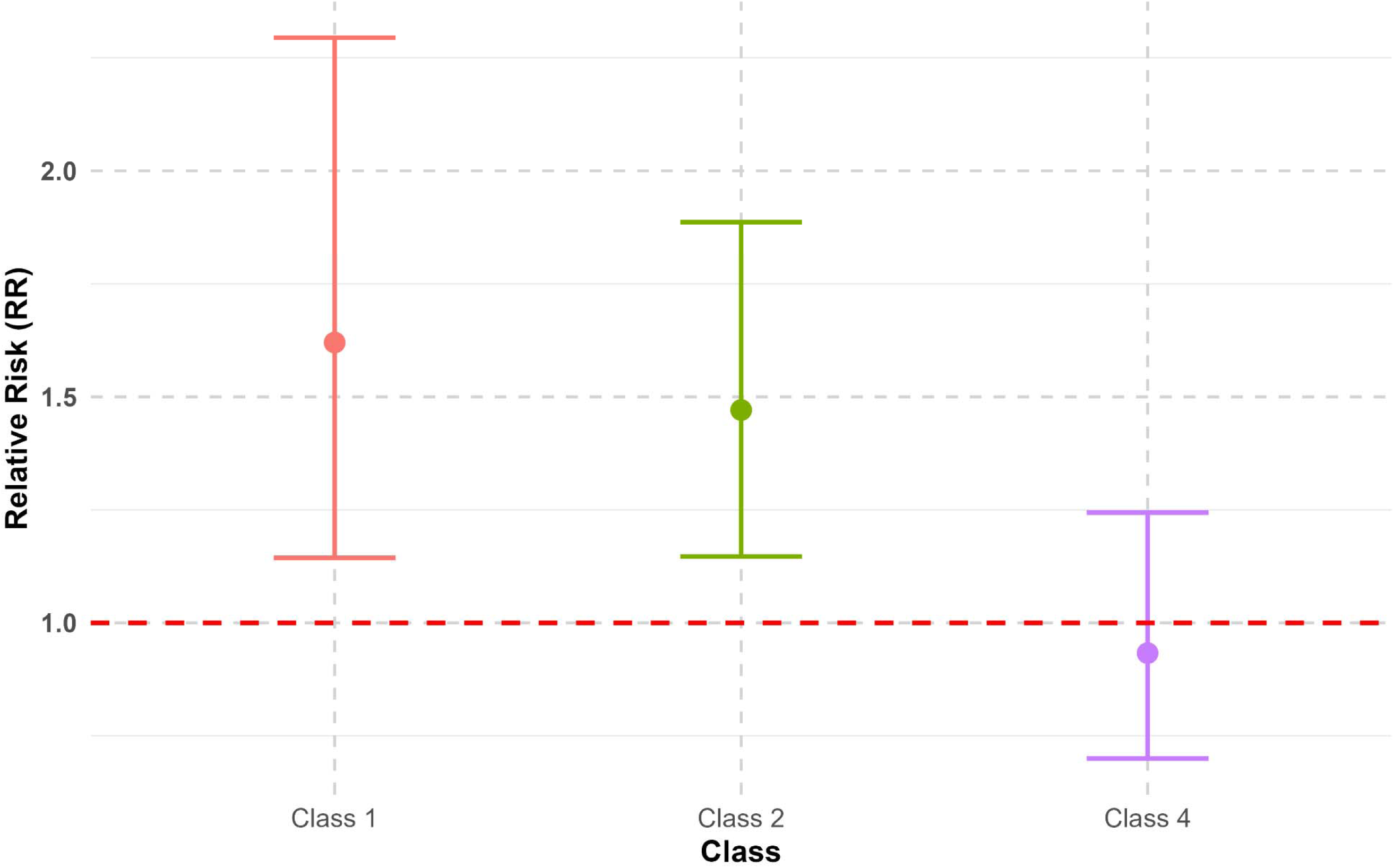
Comparison of relative risks across classes (reference: Class 3)

#### Relative Risk

To evaluate the clinical relevance of latent class membership, we estimated RR of mortality using Class 3 as the reference. Class 3 was selected due to its large sample size (n=452, 39% of the sample) and moderate clinical severity and environmental conditions. Class 1 had the highest mortality risk (RR=1.62, 95% CI: 1.14-2.29), followed by Class 2 (RR=1.47, 95% CI: 1.15=1.89). Class 4 showed the lowest mortality risk with RR=0.93 (95% CI: 0.70-1.24).

## Discussion

This study is the first to examine ICU patient profiles based on environmental factors using LCA. We retrospectively analysed data from 1,166 patients with severe COVID-19 admitted to ICUs from three hospitals. We investigated the heterogeneity in mortality by identifying subgroups, and more importantly, examined the effects of demographic, clinical, and socioeconomic covariates on mortality within these latent classes.

Our analysis revealed four distinct classes with distinct environmental profiles. Class 1 was characterised by higher temperatures and markedly lower humidity, while Class 2 showed elevated windspeed and reduced humidity. Class 3 covered the longest timespan and exhibited relatively balanced environmental conditions. In contrast, Class 4 exhibited substantial deviations, with lower temperatures and higher humidity levels. The demographic and socioeconomic suggest a relatively uniform distribution across classes from the comparisons in Table 4, these covariates therefore did not appear to be discriminators among the classes. These findings underscore the key role of environmental factors in distinguishing the latent classes.

Multivariate logistic regression analyses revealed considerable heterogeneity in mortality risk factors across the latent classes, as further supported by results from relative importance scoring and relative risk analyses. Class 1 demonstrated no significant associations with any covariates, which may be attributed to its small sample size.

Among the environmental covariates, higher relative humidity was significantly associated with reduced mortality in Class 3, the largest class in which ICU admissions encompassed all four seasons (mean relative humidity: 83.9%). Increased humidity may reduce the airborne persistence of respiratory droplets, accelerate viral decay, and support mucosal barrier function, thereby mitigating respiratory infections [16,67,68]. This association was supported by the relative importance scores, which identified humidity in Class 3 as the most influential environmental factor. These findings are consistent with previous research reporting an inverse relationship between humidity and COVID-19 mortality [31,69,70]. Of note, Romero Starke et al. [71] observed a statistically significant reduction in mortality when relative humidity exceeded a threshold of 71% which closely aligns with our findings, as the minimum recorded humidity in Class 3 was 70.9%. Temperature and windspeed were not significantly associated with mortality and consistently exhibited low relative importance scores. This lack of effect may reflect the relatively mild and stable weather conditions in the English Midlands; extreme variations in these variables were rare in our data.

Examination of demographic variables indicated that age was the only covariate significantly associated with mortality risk across Classes 2-4, with increased risk observed among older individuals, aligning with previous research [72–76]. Being of white ethnicity showed only a weak association, with a marginally significant decrease in mortality observed in Class 3. Living in a deprived area yielded no significant associations in any class, which may be due to high baseline level of deprivation in the cohort, with over half of both survivors and non-survivors originating from deprived areas.

Recent studies have found that female sex is significantly associated with reduced mortality risk [77–81]. This is posited to be due to sex-based differences in immune response, such as higher ACE2 receptor expression and greater susceptibility and severity of infection in males, stronger immune responses in females, and the protective effects of oestrogen versus the pro-inflammatory effects of testosterone. In our study, only Class 4, characterised by cold, humid conditions and gentle wind speed showed significantly lower COVID-19 mortality among females, and this was primarily observed in January and February 2021.

Clinical covariates demonstrated more consistent associations with mortality risk. Higher ICNARC scores were significantly associated with increased mortality in all classes except Class 1, reinforcing their established utility in disease severity stratification [82]. Similarly, frailty scores were significant in Class 3 and Class 4, indicating the increased vulnerability of these subgroups [83]. This importance of clinical covariates was also highlighted by the relative importance score assignment: ICNARC and frailty scores were the most influential covariates across the classes.

Numerous studies have identified a J- or U-shaped relationship between BMI and COVID-19 mortality, where underweight (BMI<18.5) and severe (Class III) obesity (BMI≥40) categories are associated with increased mortality risk compared to normal weight (BMI 19-25), overweight (BMI 25-30), and Class I obesity (BMI 30-35) categories, likely due to impaired immune function and associated comorbidities [84–91]. Interestingly, our findings supported this pattern only in Class 3 (moderate humidity), where lower BMI was significantly associated with reduced mortality risk. In this class, the interquartile range of BMI (26–34) fell within the overweight and Class I obesity range, suggesting potential physiologic effects specific to this environmental profile.

Using Class 3 (moderate humidity) as the reference group, both Class 1 and Class 2 exhibited significantly elevated RRs of mortality. Class 1 (warm, dry, windy) showed the highest RR of 1.62 (95% CI: 1.14-2.29); such environmental conditions are known to increase physiological strain through mechanisms including dehydration, impaired thermoregulation, and respiratory burden [51,92]. Class 2 (mild temperature) also showed an elevated RR of 1.47 (95% CI: 1.15-1.89), which may be partially explained by the significantly higher ICNARC score compared to Class 3.

To briefly summarise, although a substantial body of literature has examined the relationship between environmental conditions and COVID-19 mortality, our study offers a novel, nuanced perspective by exploring this association across distinct environmental profiles. By applying LCA, we captured seasonal environmental trends to reveal the associations of temperature, humidity, and wind speed with mortality risk. We found that the significance of environmental, demographic, socioeconomic, and clinical covariates, namely relative humidity, female sex, BMI, ICNARC score, frailty score, and relative humidity, emerged in a class-specific manner. These findings may have practical implications for critical care management; ICUs may benefit from integrating environmental monitoring for more nuanced risk stratification and effective resource allocation.

## Strengths and Limitations

This retrospective, multicentre cohort study used data from the first 365 days of the COVID-19 pandemic in the UK. Strengths included a low rate of missing data, which enabled robust latent class identification and analysis of heterogeneity. After class assignment, we applied multivariate logistic regression to examine variable effects on mortality across the four classes. Importantly, we found that environmental differences between classes were significantly associated with mortality risk.

Limitations include potential sampling bias. While the three hospitals represented the majority of ICU beds in Birmingham and most patients remained in the same ICU, a small number were transferred, possibly some of them with more severe illness, which may have skewed mortality outcomes.

The sample was predominantly male (67.1%), older (mean age=57.2), socioeconomically deprived (median IMD=2), and had elevated BMI (mean=30.7); additionally, the over-representation of Asian (34.3%) and White British (45.1%) patients reflects local demographics, limiting generalisability to other groups.

Another limitation could come from data collection. Frailty scores were assigned based on inpatient notes, requiring subjective clinical judgment. Misclassification may have occurred due to incomplete documentation of baseline function or comorbidities. During peak pandemic periods (April 2020 and January 2021), staffing shortages, redeployment of unfamiliar staff, and ICU overcrowding may also have negatively impacted patient prognosis.

As clinical understanding of COVID-19 evolved rapidly over this period, novel therapeutics and emerging trial results likely influenced outcomes over time. The UK vaccine rollout began in late December 2020 and by the end of February 2021, about 15% of patients admitted to the ICU aged 70 years or older were offered their first dose of vaccination. However, we lacked vaccination and treatment data for ICU patients. Future directions would include assessing how treatment and vaccination status and environmental conditions are associated with mortality, but this was beyond the scope of our study.

Finally, as this was an observational study, causal inferences cannot be made, and residual confounding, such as unmeasured comorbidities, cannot be entirely excluded. While the four-class model showed good overall fit, Class 1 included fewer patients (n=87), which may limit the precision of findings for that group. Nevertheless, the consistency in direction and significance across multiple models supports the robustness of our results.

## Conclusion

We identified four distinct subgroups defined by combinations of temperature, humidity, and windspeed, each with unique environmental risk profiles. While clinical severity and physiological vulnerability remain primary predictors of mortality, our findings suggest that environmental conditions may influence outcomes in subgroup-specific ways. This highlights the need for more nuanced risk stratification models in critical care that incorporate both patient-level and contextual factors.

Importantly, our results underscore the value of accounting for environmental variability in ICU mortality risk assessment. Integrating real-time environmental data with individual clinical profiles could enhance personalised treatment planning and optimise resource allocation in critical care.

## Data Availability

All data produced in the present study are available upon reasonable request to the authors.

## List of abbreviations

(AIC): Akaike Information Criterion
(ALCPP): Average latent class posterior probability
(BIC): Bayesian Information Criterion
(BHH): Birmingham Heartlands Hospital
(COVID-19): Coronavirus disease 2019
(GHH): Good Hope Hospital
(IMD): Index of Multiple Deprivation
(ICUs): Intensive care units
(ICNARC): Intensive Care National Audit & Research Centre
(LCA): Latent class analysis
(MWE): Medical Wire & Equipment
(OR): Odds ratios
(PICS System): Prescribing information and communication system
(QEH): Queen Elizabeth Hospital Birmingham
(RR): Relative risk
(UV): Ultraviolet
(UHB): University Hospitals Birmingham
(VLMR-LRT): Vuong-Lo-Mendell-Rubin adjusted likelihood ratio test

